# Sex and Marital-Status Differences in Delayed Initiation for Pharyngeal Cancer Treatment, Before and After Medicaid Expansion

**DOI:** 10.1101/2024.05.29.24308135

**Authors:** Jason Semprini

## Abstract

**Background:** In the United States, pharyngeal cancer has become the most common type of head and neck cancer, with 80% of cases found in males. Although disparities in treatment delays have been observed in pharyngeal patients, less is known about how policies facilitate timely care. This study aimed to estimate the association between Medicaid expansion and delaying initiation of pharyngeal cancer treatment.

**Methodology:** We extracted Surveillance, Epidemiological, End Results (SEER) case data to analyze pharyngeal cancers diagnosed between 2000-2018. The outcome of interest was a binary variable indicating if the patient initiated treatment two or more months after diagnosis. We conducted subgroup analyses by sex, marital status, and type of treatment received (surgery, radiation, chemotherapy, post-operative radiation, systemic therapy). We implement the Matrix Completion algorithm to account for staggered rollout of Medicaid expansion within our difference-in-differences design.

**Results:** Our sample included 79,433 patients diagnosed with pharynx cancer. Delayed treatment was lowest among married females receiving systemic therapy (5%), and highest among married males and females not recommended to receive surgery (43%). Generally, there was no association between Medicaid expansion and changes in delayed treatment. Subgroup analyses show that Medicaid expansion was associated with reduced treatment delays in unmarried females receiving systemic therapy (-4.5%-points), and married males receiving chemotherapy (Est. = -2.6%-points), radiotherapy (Est. = -3.1%-points), and married males not recommended to receive surgery (Est. = -4.6%-points).

**Conclusions:** Given the importance of timely pharyngeal cancer treatment, health systems must identify and address the drivers of treatment delays to advance cancer equity.

## Introduction

In the United States, pharyngeal cancer has emerged as the most common type of new head and neck cancer diagnosis^1^. An overwhelming majority (83%) of these pharyngeal cancers are diagnosed in men, underscoring the need for expanding prevention and control efforts for males across the country^2–4^. Although pharyngeal cancer carries a more favorable outlook than other cancers within the head and neck^5^, the rising incidence of these often late-stage pharyngeal cancer diagnoses threatens cancer control systems^6^. Late-stage incidence is growing and again, there were considerable disparities by sex, as localized diagnoses comprised 19% of all female cases and just 11% of all male cases^2,6^. These trends may exacerbate existing disparities and increase the disproportionate burden of pharyngeal cancer experienced by thousands of males in the United States.

Advancements in treatment modalities have improved survival rates for oropharyngeal cancer. However, these advancements rely heavily on access to high-quality, *timely* care^7^. Prior work has illuminated that disparities in pharyngeal cancer survival may be due to disparities in care quality^8^. Although timing has proven critical for survival^9^, research has illuminated the persisting disparities in delayed treatment by race, socioeconomic status, and geography^10–12^. Still, questions remain.

The existing evidence illuminating the importance of and disparities in receiving prompt treatment has focused on head and neck cancers, generally, and less focused on pharyngeal cancer, specifically. Moreover, only one study attempted to explore sex-based differences in pharyngeal cancer delays and found no differences between males and females^13^. However, this study focused on surgery and may not generalize to other types of pharyngeal cancer treatment modalities. Finally, while the literature has aimed to illuminate pharyngeal cancer disparities, few studies have explored potential policies to address them.

One policy, the Affordable Care Act (ACA)’s Medicaid Expansion has, in fact, been heavily studied for its impact on cancer outcomes across the care continuum^14,15^. Research showed that the ACA’s Medicaid Expansion was associated with reduced treatment initiation delays for patients with oral cavity cancer, but possibly associated with greater treatment delays for patients with HPVa pharyngeal cancer^16^. The decline in treatment delays following Medicaid Expansion for oral cavity patients could be due to improved access to Medicaid dental coverage and earlier detection^17^. Dental services and earlier detection, however, are less likely mechanisms for explaining the potential relationship between Medicaid Expansion and earlier treatment of pharyngeal cancer.

Other explanations are more likely. One possibility is that Medicaid Expansion increased access to care and, therefore, improved time-to-treatment measures by removing barriers to care for uninsured or underinsured adults with pharyngeal cancer. This explanation, however, could only improve access for those gaining Medicaid through expansion and would not necessarily improve outcomes for adults with commercial or Medicare insurance. Although lower socioeconomic status subgroups would benefit the most from gaining insurance, Medicaid Expansion would, at most, reduce uninsurance by merely 2-5% overall^14,18,19^. Alternatively, Medicaid expansion could increase delays in treatment initiation if health systems delayed care because their capacity to deliver services could not meet the influx of new patients gaining access to the healthcare system. This alternative explanation would affect all patients, not just those gaining Medicaid. Unfortunately, Medicaid expansion literature rarely considers broad, system-level perspectives in lieu of the narrow perspective of patients expected to benefit from gaining Medicaid after expansion. This limited perspective has hindered our understanding of how major health policies impacted access to care differently across the entire population, with major implications for our pursuit of cancer equity^20^.

### Objective

This study aimed to evaluate the effect of Medicaid Expansion on treatment initiation delays for patients diagnosed with pharyngeal cancer. In addition to estimating effects for a variety of treatment modalities and exploring baseline disparities, this study also tests if the effects of expansion significantly vary by sex and marital status; a proxy for socioeconomic status and social support.

## Materials and Methods

### Data

We analyzed specialized Head and Neck Cancer data from the Surveillance, Epidemiological, End Results (SEER) program^21^. This specialized datafile includes an HPV recode collected by SEER and assigned to type of the following cancers based on the CS Collaborative Stage Scheme: Hypopharynx, Nasopharynx, Oropharynx, Pharyngeal Tonsil, Pharynx Other, Palate Soft, Tongue Base^22^.

### Inclusion Criteria

The analytic datafile includes all cases of pharyngeal cancers diagnosed between 2000-2018, among the following states (California, Connecticut, Georgia, Hawaii, Iowa, Kentucky, Louisiana, New Jersey, New Mexico, Utah) and registries (Detroit, Seattle). Cases were restricted to first-primary, malignant tumors. Cancers diagnosed on an autopsy or death certificate only were excluded.

### Variables

Our outcome is a binary variable indicating if the patient initiated cancer treatment 2 or more months after diagnosis. This variable was derived from SEER’s months to treatment initiation variable, which calculates the number of calendar months that have passed since the month of diagnosis. Although limited, this is the only available measure of time-to-treatment within the public SEER datafile^21^.

SEER also includes the year of diagnosis and state of residence, which we leverage for our identification strategy (see design below). We also utilize independent variables found in SEER which measure individual factors (age, sex, race, ethnicity, marital status) and tumor characteristics (stage of diagnosis, site schema, histology, grade). Each individual and tumor measures were modelled as mutually exclusive categories of binary control variables in all analyses. We also used sex (male, female) and marital status (married, unmarried) to stratify the sample. SEER also includes data on the first sequence of treatment (primary surgery, lymph nodes removed, radiation, chemotherapy, surgery not recommended, post-operative radiation, systemic therapy). These treatment measures are not necessarily mutually exclusive, so we analyze time-to-treatment initiation trends for each of these types of treatment measures.

Missing HPV data was imputed through Multiple Imputation Chained Equations^23^ (50x) Logistic Regressions with histology, grade, schema as predictors^17,24,25^. Given that the level of missingness varies over time (∼70% in 2010 and 20% in 2017)^22^, and high proportion of estimated HPVa OPC (>70%), we do not restrict the sample to HPV positive cases but instead add imputed HPV codes to all models.

### Exposure

The exposure of interest is Medicaid expansions under the ACA. Although most prior ACA policy evaluations modelled expansion at a single time period (2014+), in reality some states expanded their Medicaid program up to ACA levels (>=138% FPL) before and after^26,27^. Two participating SEER states (California and Washington) expanded their Medicaid income eligibility guidelines by 2011^26^. Additionally, another SEER state (Louisiana) expanded Medicaid by 2016^27^. The other SEER expansion states (Connecticut, Hawaii, Iowa, Kentucky, Michigan, New Jersey, New Mexico) expanded in 2014^27^. Only two SEER states (Georgia, Utah), did not expand Medicaid before 2018^27^. Amidst this staggered exposure to Medicaid expansion, all patients will be considered exposed to expansion if they resided in an expansion state during or after that state’s expansion year.

### Design

Our goal is to estimate the association between Medicaid expansion and changes in the probability of delaying treatment 2 or more months from diagnosis. To estimate a causal effect of this policy, we construct a Difference-in-Differences design to leverage the staggered rollout of expansion by states, over time^28,29^. By including state and year fixed-effects, our design accounts for time-invariant differences in delayed treatment patterns between states, and temporal trends in treatment delays affecting all states. The resulting association can be interpreted causally under the assumption that there no unobserved variables contributing to differences in treatment delay trends between expansion and non-expansion states. Put differently, under the parallel trends assumption, we assume that in the absence of Medicaid expansion, treatment delay trends would have remained parallel over time for both expansion and non-expansion states^30^. Although this assumption cannot be empirically proved, we test the validity of this assumption by constructing differential pre-trend tests under an event-history study framework^31^.

Additionally, recent applied econometric work has highlighted the potential threats of Difference-in-Differences designs under staggered rollout^32,33^. To overcome these threats, namely related to negative weights and dynamic treatment effects, we implement an emerging econometric approach: Matrix Completion^34^. In our study, this approach constructs parallel, counterfactual trends of states which have not yet expanded to create valid comparison groups between expansion states and states which have yet to (or never) expand Medicaid^34^. The resulting estimate aggregates all comparisons of treatment delay trends between expansion and not yet (or never) expansion states into an average treatment effect estimate.

### Statistical Analysis

The probability of delaying treatment 2 or more months from diagnosis is a binary outcome and analyzed by a linear probability regression model. For inference, standard errors were clustered at the state-level^35^. All analyses were conducted in STATA v. 18, using the Matrix Completion package^36^.

## Results

### Summary Statistics

The analytic sample included 79,433 cases (unique patients diagnosed with pharyngeal cancer) between 2000-2018 (Table 1). 68.4% of all cases were attributed to HPV, a result consistent between the imputed and raw codes derived from SEER. Only 11% of cases were diagnosed at localized stages, and 20% of cases were diagnosed at distant stages. Males constituted an overwhelming majority of the cases (80.2%). 7.4% of the cases were Hispanic and 10.9% of the cases were non-Hispanic Black patients. 53.3% of the cases were married. Table 1 also reports the proportion of cases receiving specific treatment recommendations. The highest proportion of treatment received was radiation (79.6%) and the lowest proportion of treatment received was Systemic Therapy (22.3%).

**Table 1:** Sample Summary Characteristics (2000-2018)

|  | N | (%) |
| --- | --- | --- |
| ***Tumor Characteristics*** |  |  |
| Squamous Cell Carcinoma | 72,681 | (91.5) |
| Tongue | 23,194 | (29.2) |
| Tonsil | 27,563 | (34.7) |
| Other Pharyngeal | 28,675 | (36.1) |
| Localized Stage | 8,817 | (11.1) |
| Regional Stage | 51,393 | (64.7) |
| Distant Stage | 15,966 | (20.1) |
| Unknown Stage | 3,225 | (4.1) |
| ***Patient Characteristics*** |  |  |
| Male | 63,705 | (80.2) |
| <40 | 2,693 | (3.4) |
| 40-49 | 3,289 | (4.1) |
| 45-49 | 7,451 | (9.4) |
| 50-54 | 12,074 | (15.2) |
| 55-59 | 14,695 | (18.5) |
| 60-64 | 13,583 | (17.1) |
| 65-69 | 10,406 | (13.1) |
| 70+ | 15,251 | (19.2) |
| Hispanic | 5,838 | (7.4) |
| non-Hispanic Black | 8,658 | (10.9) |
| non-Hispanic White | 58,304 | (73.4) |
| Married | 42,338 | (53.3) |
| ***Treatment Characteristics*** |  |  |
| Surgery - Primary | 23,036 | (29.0) |
| Surgery - Any | 24,386 | (30.7) |
| Surgery NOT recommended | 49,010 | (61.7) |
| Lymph Nodes Removed | 28,596 | (36.0) |
| Chemotherapy | 50,837 | (64.0) |
| Radiation | 63,229 | (79.6) |
| Combination Radiation | 29,311 | (36.9) |
| Systemic Therapy | 17,714 | (22.3) |
| ***Months from Dx to Tx*** |  |  |
| 0 months | 27,563 | (34.7) |
| 1 month | 26,991 | (34.0) |
| 2 months | 12,741 | (16.0) |
| 3+ months | 12,106 | (15.2) |
| **Total** | **79,433** |  |

Table 1 reports the summary statistics of the analytic sample. Sample includes all pharyngeal cases within the Specialized SEER-HPV HNC datafile (2000-2018).

Overall, 68.7% of the cases initiated treatment within 1 month of diagnosis (Table 1). to 2010, baseline rates of delayed treatment initiation (2 or more months from diagnosis) were 35.1% (Table 2). These rates are statistically significantly different (based on confidence interval estimates) across types of treatment modalities (Supplemental Exhibit 1). The lowest rate of delayed treatment is found in cases receiving systemic therapy (9.6%). The highest rate of delayed treatment is found in cases not recommended to receive surgery (37.7%). When examining the rate of delayed treatment by sex and marital status, we see little evidence of baseline differences by sex (Table 2, Figure 1). Rather, we see much larger differences by marital status.

**Table 2.** Baseline (2000-2010) Rates of Delaying Treatment 2+ Mo.

|  | **Both Sexes** | **Females** | **Males** |
| --- | --- | --- | --- |
| ***Married & Unmarried*** |  |  |  |
| All | 0.351 | 0.360 | 0.348 |
| Surgery – Primary Site | 0.109 | 0.119 | 0.106 |
| Lymph Nodes Removed | 0.121 | 0.126 | 0.119 |
| Chemotherapy | 0.246 | 0.242 | 0.248 |
| Radiation | 0.243 | 0.245 | 0.243 |
| Surgery NOT Recommended | 0.377 | 0.398 | 0.372 |
| Combination Radiation | 0.106 | 0.104 | 0.107 |
| Systemic Therapy | 0.096 | 0.088 | 0.098 |
| ***Not Married*** |  |  |  |
| All | 0.363 | 0.362 | 0.364 |
| Surgery – Primary Site | 0.126 | 0.135 | 0.122 |
| Lymph Nodes Removed | 0.135 | 0.147 | 0.132 |
| Chemotherapy | 0.285 | 0.265 | 0.291 |
| Radiation | 0.285 | 0.264 | 0.292 |
| Surgery NOT Recommended | 0.435 | 0.434 | 0.435 |
| Combination Radiation | 0.121 | 0.120 | 0.121 |
| Systemic Therapy | 0.108 | 0.124 | 0.103 |
| ***Married*** |  |  |  |
| All | 0.252 | 0.257 | 0.251 |
| Surgery – Primary Site | 0.097 | 0.103 | 0.096 |
| Lymph Nodes Removed | 0.111 | 0.104 | 0.113 |
| Chemotherapy | 0.218 | 0.215 | 0.218 |
| Radiation | 0.211 | 0.220 | 0.210 |
| Surgery NOT Recommended | 0.324 | 0.341 | 0.321 |
| Combination Radiation | 0.097 | 0.087 | 0.099 |
| Systemic Therapy | 0.089 | 0.046 | 0.095 |

Table 2 reports the baseline rates (proportion) of cases delaying treatment initiation 2 or more months from diagnosis. Proportions are stratified by sex and marital status, for specific treatment modalities received. Proportions are on a 0-1 binary scale.

**Figure 1:**
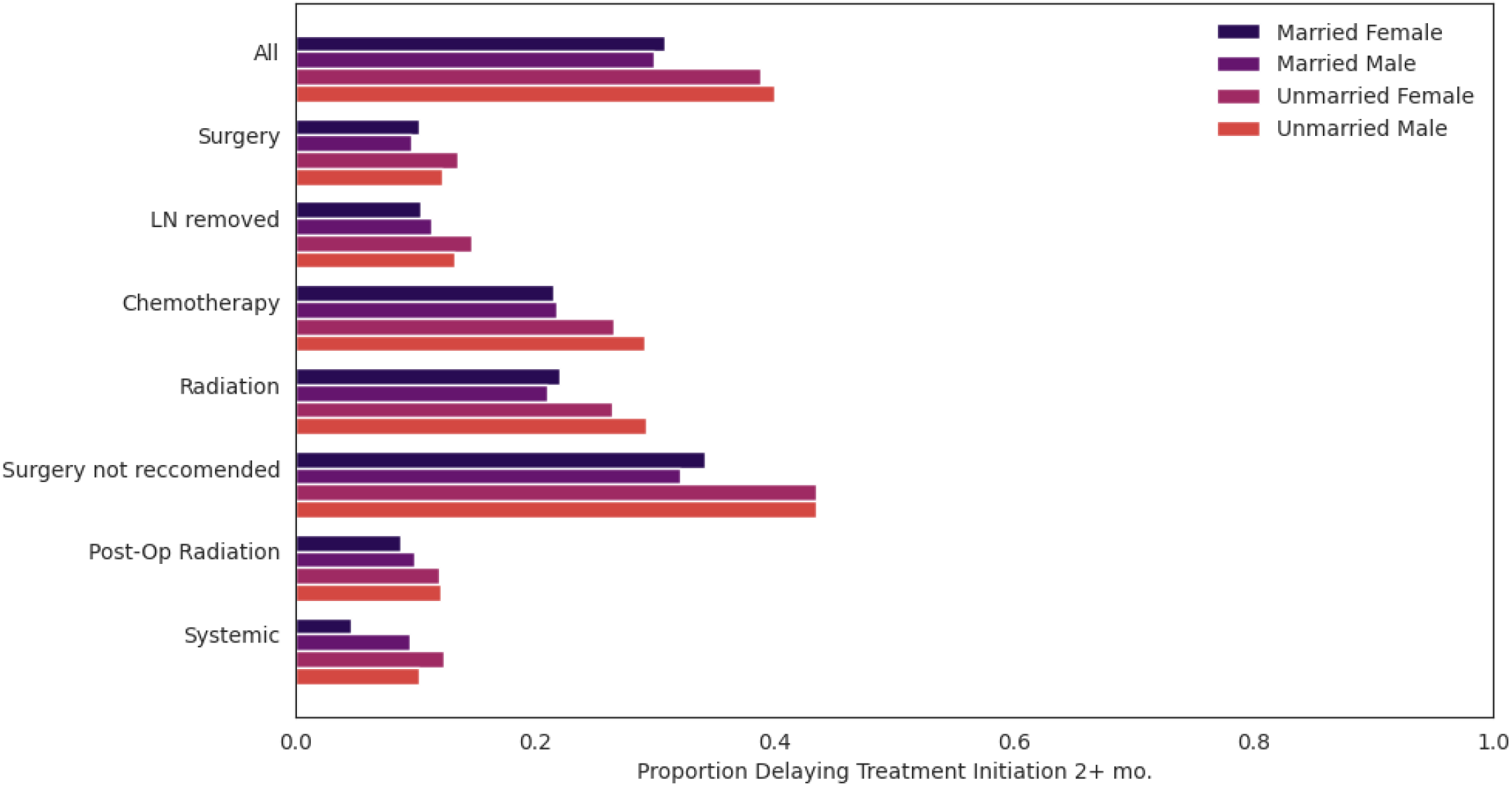
Baseline Rates of Treatment Delay (2 or more months), by sex and marital status. Figure 1 shows the baseline (2000-2010) rates (proportion) of cases delaying treatment initiation two or more months from diagnosis. Rates are stratified by sex, marital status, and treatment modality.

Overall, and within each sex, delayed treatment rates are higher among unmarried than married cases. However, we do observe heterogeneity by sex and marital status across treatment modalities. The lowest rate of delayed treatment is found in married females receiving systemic therapy (4.6%), which is significantly higher in both married males (9.5%) and unmarried females (12.4%). The highest rate of delayed treatment is found among unmarried females (43.4%) and unmarried males (43.5%) not recommended to receive surgery. These differences are not statistically different from each other but are statistically different (∼10%-points higher) than rates of married counterparts. Figures 2-3 visualize these trends in treatment delays over time, by sex and marital status, showing that rate of treatment delays are generally rising over time.

**Figure 2:**
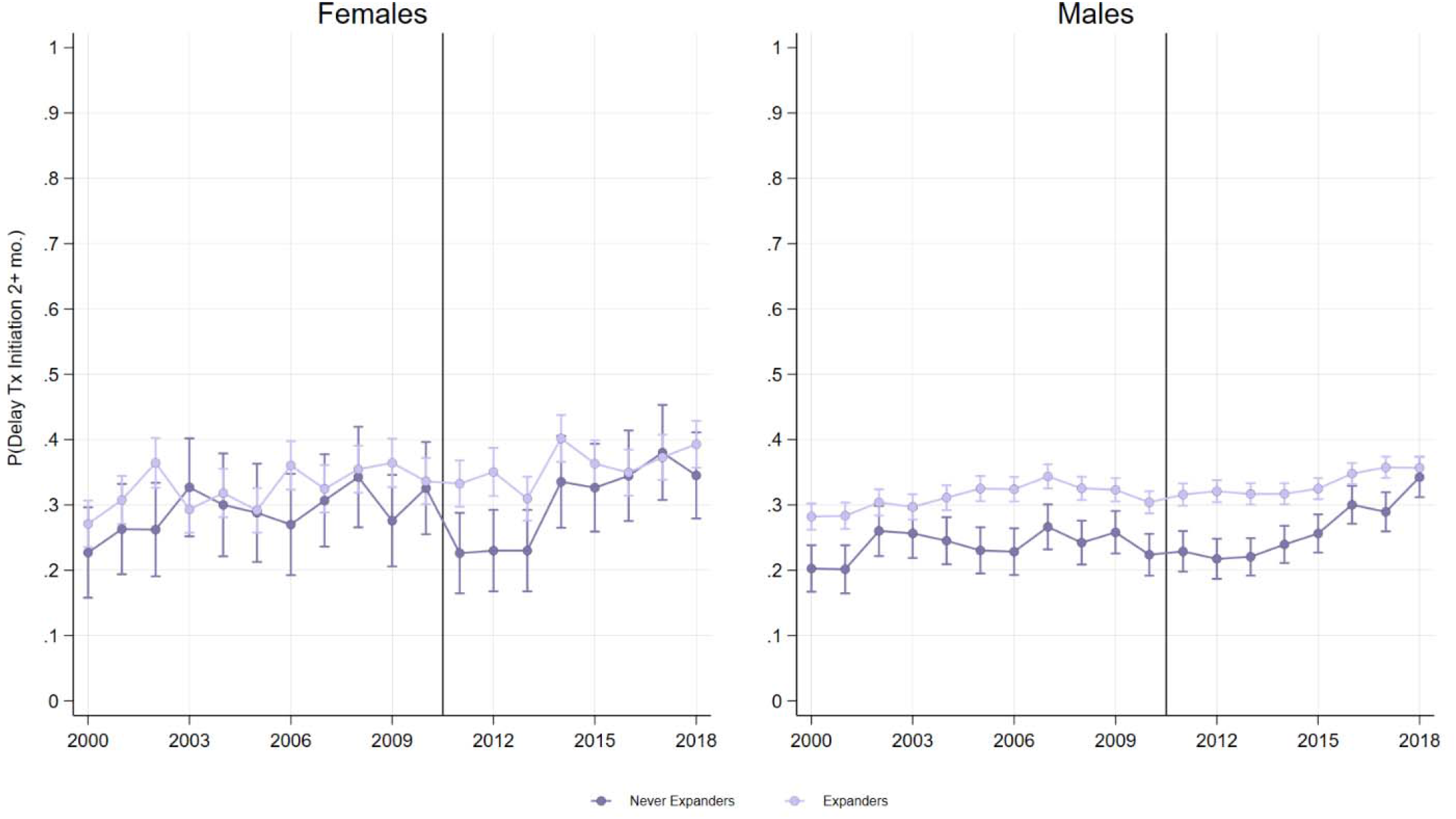
Trends in Treatment Delays (2000-2018), by sex. Figure 2 shows the proportion of cases delaying treatment initiation two or more months from diagnosis, by sex. Solid vertical line indicates year 2010, when Medicaid expansion began under the ACA in some states.

**Figure 3:**
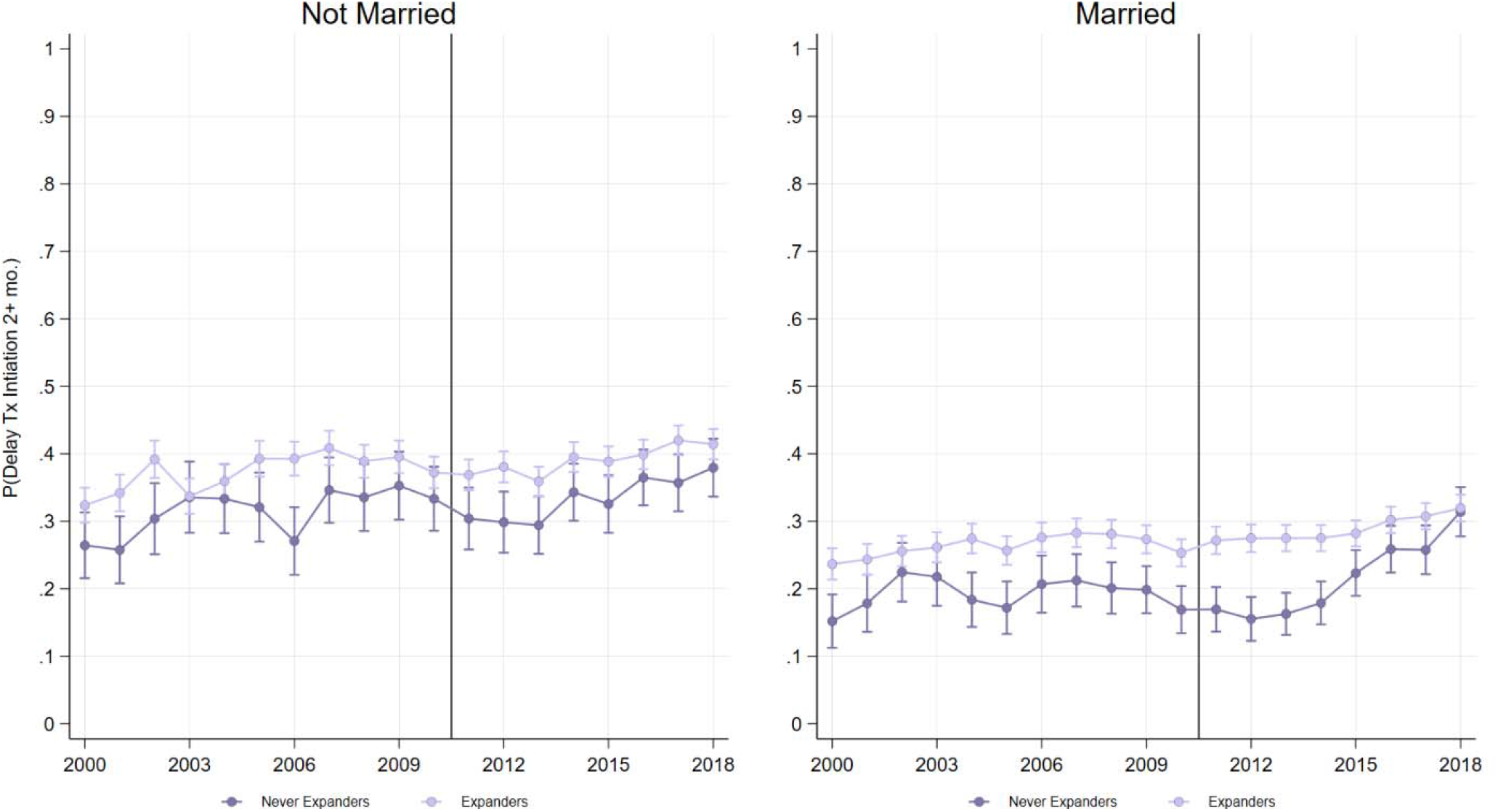
Trends in Treatment Delays (2000-2018), by marital status /. Figure 2 shows the proportion of cases delaying treatment initiation two or more months from diagnosis, by marital status. Solid vertical line indicates year 2010, when Medicaid expansion began under the ACA in some states.

### Effects of Medicaid Expansion on Delayed Treatment

Overall, there was no association between Medicaid expansion and treatment delays (Table 3). Similarly, there was no association between Medicaid expansion and treatment delays for any type of treatment modality. When stratifying by sex, we see no evidence that Medicaid expansion was associated with treatment delays in either females or males. In married males and unmarried females, we found no statistically significant associations between Medicaid expansion and treatment delays.

**Table 3:**
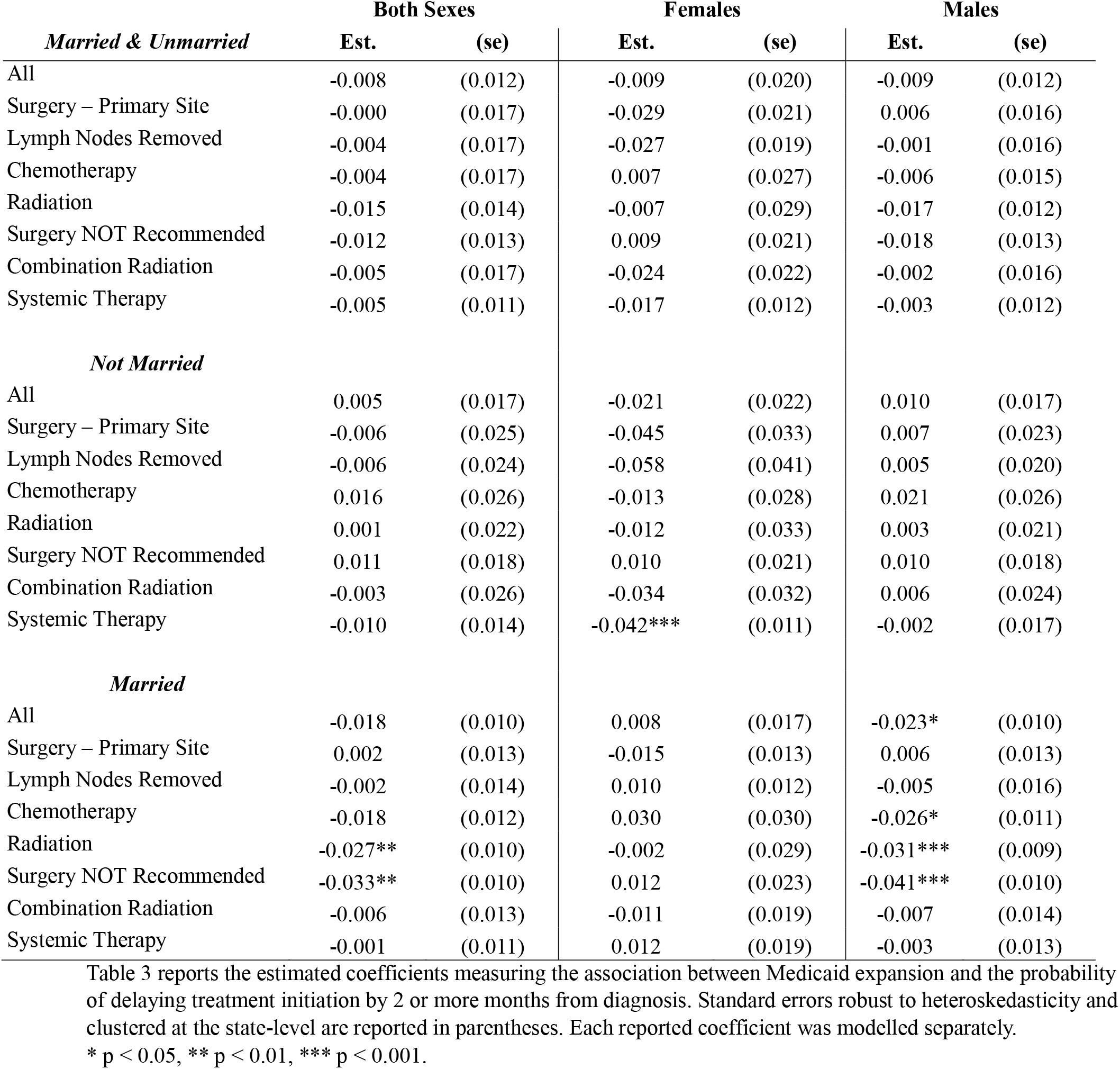
Estimated association between Medicaid expansion and delaying treatment initiation 2+ months.

|  | **Both Sexes** |  | **Females** |  | **Males** |  |
| --- | --- | --- | --- | --- | --- | --- |
| *Married & Unmarried* | **Est.** | **(se)** | **Est.** | **(se)** | **Est.** | **(se)** |
| All | -0.008 | (0.012) | -0.009 | (0.020) | -0.009 | (0.012) |
| Surgery – Primary Site | -0.000 | (0.017) | -0.029 | (0.021) | 0.006 | (0.016) |
| Lymph Nodes Removed | -0.004 | (0.017) | -0.027 | (0.019) | -0.001 | (0.016) |
| Chemotherapy | -0.004 | (0.017) | 0.007 | (0.027) | -0.006 | (0.015) |
| Radiation | -0.015 | (0.014) | -0.007 | (0.029) | -0.017 | (0.012) |
| Surgery NOT Recommended | -0.012 | (0.013) | 0.009 | (0.021) | -0.018 | (0.013) |
| Combination Radiation | -0.005 | (0.017) | -0.024 | (0.022) | -0.002 | (0.016) |
| Systemic Therapy | -0.005 | (0.011) | -0.017 | (0.012) | -0.003 | (0.012) |
| *Not Married* |  |  |  |  |  |  |
| All | 0.005 | (0.017) | -0.021 | (0.022) | 0.010 | (0.017) |
| Surgery – Primary Site | -0.006 | (0.025) | -0.045 | (0.033) | 0.007 | (0.023) |
| Lymph Nodes Removed | -0.006 | (0.024) | -0.058 | (0.041) | 0.005 | (0.020) |
| Chemotherapy | 0.016 | (0.026) | -0.013 | (0.028) | 0.021 | (0.026) |
| Radiation | 0.001 | (0.022) | -0.012 | (0.033) | 0.003 | (0.021) |
| Surgery NOT Recommended | 0.011 | (0.018) | 0.010 | (0.021) | 0.010 | (0.018) |
| Combination Radiation | -0.003 | (0.026) | -0.034 | (0.032) | 0.006 | (0.024) |
| Systemic Therapy | -0.010 | (0.014) | -0.042*** | (0.011) | -0.002 | (0.017) |
| *Married* |  |  |  |  |  |  |
| All | -0.018 | (0.010) | 0.008 | (0.017) | -0.023* | (0.010) |
| Surgery – Primary Site | 0.002 | (0.013) | -0.015 | (0.013) | 0.006 | (0.013) |
| Lymph Nodes Removed | -0.002 | (0.014) | 0.010 | (0.012) | -0.005 | (0.016) |
| Chemotherapy | -0.018 | (0.012) | 0.030 | (0.030) | -0.026* | (0.011) |
| Radiation | -0.027** | (0.010) | -0.002 | (0.029) | -0.031*** | (0.009) |
| Surgery NOT Recommended | -0.033** | (0.010) | 0.012 | (0.023) | -0.041*** | (0.010) |
| Combination Radiation | -0.006 | (0.013) | -0.011 | (0.019) | -0.007 | (0.014) |
| Systemic Therapy | -0.001 | (0.011) | 0.012 | (0.019) | -0.003 | (0.013) |

Table 3 reports the estimated coefficients measuring the association between Medicaid expansion and the probability of delaying treatment initiation by 2 or more months from diagnosis. Standard errors robust to heteroskedasticity and clustered at the state-level are reported in parentheses. Each reported coefficient was modelled separately.
\* p < 0.05, \*\* p < 0.01, \*\*\* p < 0.001.

When stratifying by sex and marital status, however, we begin to observe some evidence that Medicaid expansion differentially affected treatment delays. As mentioned above, baseline rates of treatment delays were lowest among married females receiving systemic therapy. We found, however, that Medicaid expansion was associated with a 4.2%-point reduction (p < 0.001) in the probability of delaying treatment initiation in unmarried females receiving systemic therapy. This association reflects a 34% reduction relative to baseline rates. The estimate in this group of unmarried females was statistically and significantly different than the respective estimate in other groups receiving systemic therapy (Table 3).

For married males, Medicaid expansion was associated with a 2.3%-point reduction (p < 0.05), in delaying treatment initiation two months or more, reflecting a 9% reduction from baseline rates. The association was significant for unmarried males undergoing chemotherapy (Est. = -0.026, se = .011, p < 0.05) and radiotherapy (Est. = -0.031, se = 0.009, p < 0.001). These estimates reflect a 12% and 15% relative change from baseline, respectively. The largest estimate in unmarried males was found in those not recommended to receive surgery, where Medicaid expansion was associated with a 4.1%-point reduction (p < 0.001) in the probability of delaying treatment two or more months. This estimate reflected a 15% relative change from baseline. The associations between Medicaid expansion and delayed treatment for unmarried males receiving radiation and those not recommended to receive surgery were statistically different than the respective association among counterpart groups.

The event-history study specification supports the validity of our estimates, as none of the models with statistically significant associations between Medicaid expansion and delayed treatment included statistically significant differential trends between expansion and non-expansion states in the two years leading up to expansion (Supplemental Exhibit 2).

## Discussion

This study examined treatment initiation delays for patients with pharyngeal cancer. Before 2010, initiation delays varied by type of treatment received, with less heterogeneity by sex than by marital status. After 2010, we observe that treatment delays increased broadly, with little evidence that Medicaid expansion contributed to the rise in delayed treatment. However, there appeared to be some evidence that Medicaid expansion reduced treatment delays in specific subgroups (i.e., unmarried females receiving systemic therapy, married males receiving chemotherapy or radiotherapy). Such heterogeneity has implications for health equity. Considering marital status as a proxy for socioeconomic status or social support, prior to 2010 unmarried females had higher rates of treatment delays than married females. This disparity was especially pronounced for systemic therapy. Although disparities persist beyond 2010, Medicaid expansion may have mitigated socioeconomic disparities in treatment delays among females receiving systemic therapy. However, because there was no such association in unmarried males, Medicaid expansion may have intensified sex-based disparities among unmarried populations receiving systemic therapy for pharyngeal cancer. In contrast, Medicaid expansion reduced the proportion of married males delaying treatment initiation for chemotherapy and radiation therapy. Since, however, treatment delays in this population were already lower than unmarried counterparts prior at baseline, Medicaid expansion may have widened socioeconomic disparities within males while mitigating sex-based disparities.

The dynamic trends in treatment initiation delays for pharyngeal cancer patients revealed a complex relationship between types of treatment received and patient factors such as state of residence, sex, and marital status. The rising, heterogenous rates of delayed treatment initiation underscore the need for targeted interventions that address the specific needs of subgroups, while fitting the context and constraints of health systems. Future research should evaluate how policies and programs providing social (i.e., navigation) and financial support could reverse the rising trends in treatment delays. Additionally, further research is needed to examine the long-term impact of treatment delays on patient-centered outcomes such as survival and quality of life. This evidence could inform clinical decision-making and guide the development of evidence-based guidelines for the management of pharyngeal cancer.

### Limitations

SEER registries cover approximately 34% of the U.S. population and comprise under 50% of all cancer diagnoses. For these reasons, the extent to which our results generalize to the U.S. population may be limited. Moreover, SEER does not track measures relevant for assessing socioeconomic disparities (i.e., income, education, employment) or reasons why treatment was delayed (i.e., insurance status). We use marital status, but that variable could be confounded by many factors. While can definitively say is that treatment delays varied by marital status, we cannot explain the causes of such variation. Regarding our research design, all quasi-experimental observational studies are subject to potential bias from unobserved confounding. We take steps to minimize the threat of this potential bias by implement a novel, yet valid conceptually intuitive, method to support our internal validity. We also assess the validity of our research design and identification strategy by reporting the results of an event-history study. Still, we urge readers to view our results without causal interpretations.

### Conclusions

As the incidence of pharyngeal cancer continues to rise in the United States, access to high-quality treatment remains paramount for reversing sociodemographic disparities in patient outcomes. High-quality treatment typically requires *timely* treatment. The extent to which treatment delays persist or contribute to pharyngeal cancer disparities, has not been well understood. Using population-based data, we first show that rates of delaying treatment initiation for pharyngeal cancer varied across a variety of treatment modalities. We also show that, prior to 2010, treatment delays were generally consistent between males and females, but were much higher in unmarried compared to married patients. We also showed that after 2010, the proportion of pharyngeal cases delaying treatment two or more months from diagnosis has increased. However, there is little evidence suggesting this was due to Medicaid expansion. Only in a few subgroups (unmarried females receiving systemic therapy, married males receiving chemotherapy or radiotherapy) did we estimate an association between Medicaid expansion and reduced treatment delays. How the dynamic, heterogeneous trends in delaying pharyngeal cancer treatment will impact socioeconomic disparities in mortality and quality of life remains unknown. Research investigating the causes and, more importantly, potential health system policies addressing treatment delays could advance health equity as the burden of this disease continues to grow.

## Supporting information

Supplemental Exhibits 1-2

## Acknowledgments

None.

## Data Availability

Restricted by third-party (NCI). SEER case files and analytic STATA code are available on the corresponding author’s public repository [BLIND DURING PEER REVIEW].

